# Sex Differences in Vulnerability to Tau Pathology: Impact on Cognitive Decline

**DOI:** 10.1101/2025.05.25.25328324

**Authors:** Yashar Zeighami, Cecilia Tremblay, Mahsa Dadar

## Abstract

**Introduction:** While the link between presence of amyloid and tau pathologies, neurodegeneration, and cognitive decline in aging individuals is established, it is less clear whether there are sex differences in vulnerability to these pathologies.

**Methods:** 1,464 participants (7,168 longitudinal assessments, 4.77±3.78 years of follow-up) were included from the National Alzheimer’s Coordinating Center (NACC) database. Longitudinal mixed effects and mediation models examined the sex differences across cognitive decline trajectories of amyloid (A), tau (T), and neurodegeneration (N) groups.

**Results:** A^+^T^-^ males showed faster cognitive decline compared to A^+^T^-^ females (p<0.005), whereas A^+^T^+^ females showed steeper cognitive decline compared to A^+^T^+^ males (p<0.0001). Sex also marginally moderated the mediating effect of tau on the relationship between amyloid and cognitive decline (p=0.046).

**Discussion:** Sex differences in vulnerability to tau pathology in presence of amyloid can shape cognitive decline trajectories.

**Highlights:**

- A^+^ T^-^ males showed faster cognitive decline compared to A^+^ T^-^ females
- A^+^ T^+^ females showed faster cognitive decline compared to A^+^ T^+^ males
- Tau status significantly mediated the relationship between amyloid status and cognitive decline
- Sex marginally moderated the mediating relationship between amyloid, tau, and cognitive decline
- The findings point to sex differences in the impact of tau pathology on cognition

## Introduction

Alzheimer’s disease (AD) is the most common neurodegenerative disease and cause of dementia in the aging population. Based on the definition of the National Institute on Aging - Alzheimer’s Association (NIA-AA) Alzheimer’s diagnostic framework to biologically define AD, AD is characterized by the presence of three distinct biomarkers, amyloid beta deposition (A), pathological tau (T), and neurodegeneration (N)^1,2^. Clinically, positivity on amyloid and tau markers can be established through amyloid and tau Positron Emission Tomography (PET) or cerebrospinal fluid (CSF) assessments, whereas neurodegeneration can be determined based on magnetic resonance imaging (MRI) or Fludeoxyglucose (FDG) PET findings^2^. Longitudinal trajectories of AD amyloid-tau-neurodegeneration (ATN) biomarkers have been assessed in elderly persons, with the most common pattern being the amyloid pathology biomarker that emerges first^2^. Specifically, based on these, individuals that are negative on all three markers (A^-^ T^-^ N^-^) are considered as normal or without AD pathology, individuals that are positive on amyloid and negative on tau and neurodegeneration (A^+^ T^-^ N^-^) are considered to be at earlier stages of the disease, and individuals that are positive on amyloid and tau (A^+^ T^+^ N^-^ or A^+^ T^+^ N^+^) are considered as AD (earlier or later stages, respectively). Finally, individuals that are negative on amyloid but positive on tau or neurodegeneration (A^-^ T^+^ N^-^, A^-^ T^-^ N^+^, or A^-^ T^+^ N^+^) are considered to have non-AD pathologies, and individuals that are positive on amyloid and neurodegeneration but negative on tau are considered as having AD pathology as well as other non-AD pathology (A^+^T^-^N^+^) that contributes to neurodegeneration^2,3^.

A growing body of research on sex differences in AD points to significantly higher prevalence and burden of tau pathology and cognitive deficits in females compared to males^4–7^. Sex-specific differences in pathology burden might also have implications for disease management and treatment, as sex differences in treatment efficiency have been recently reported^8,9^. Finally, while previous studies suggest a mediating effect of tau pathology on the associations between amyloid burden and cognitive decline^10–13^, it is unclear whether these associations differ between sexes. As such, given the accumulating literature pointing to sex differences in AD pathology burden and cognitive decline trajectories^3,14–18^, more studies are needed to disentangle the effects of sex on the associations between amyloid and tau biomarkers and cognitive decline.

This study aimed to assess the sex differences in longitudinal cognitive trajectories, specifically disentangling the contribution of amyloid, tau, and neurodegeneration, by comparing ATN biomarker-based groups using a large cohort of aged individuals along the AD continuum, from non-demented to AD dementia. To do so, we took advantage of the large number of participants with amyloid, tau, and neurodegeneration biomarker status information as well as long term longitudinal cognitive assessments available from the National Alzheimer’s Coordinating Center (NACC)^19,20^ database. We hypothesized amyloid and tau positive biomarkers groups with neurodegeneration to show a steeper cognitive decline followed by groups positive for amyloid without neurodegeneration or tau pathology and expected to find a steeper cognitive decline in females, especially in groups with tau positive biomarkers.

## Methods

Data was obtained from the National Alzheimer’s Coordinating Center (NACC, https://naccdata.org/; September 2024 data freeze) database^19–22^. Ethics approval was obtained from the participants by the local institutional review board from each National Institute on Aging Alzheimer’s Disease Centers (ADRCs) which contributed the data to NACC. Participants were included if they had demographics information as well as PET or CSF based amyloid and tau status (using variables AMYLPET [N = 1074], AMYLCSF [N = 390], TAUPETAD [N = 886], and CSFTAU [N = 578]) and visual assessment of hippocampal atrophy (using the variable HIPPATR [N = 1464]) available. These binary values reflecting abnormality on amyloid and tau, and hippocampal atrophy were used to derive ATN classification for all participants. Clinical dementia rating sum of boxes (CDR-SB) scores were used as a measure of global cognition (using the variable CDRSUM). Exclusion criteria included missing ATN information, clinical diagnosis of non-AD neurological disorders (e.g. Lewy body dementia, frontotemporal dementia, etc.) based on the NACC variable NACCETPR, participants with ages below 55 or above 95, and participants without longitudinal follow up assessments including CDR-SB information available. Furthermore, due to the small number of participants with A^-^ T^+^ N^+^ (2 females and 12 males), this group was not included in the analyses. This resulted in a total of 1464 participants with 7168 longitudinal timepoints included (average follow-up: 4.77 ± 3.78 years, 4.9 ± 3.2 visits). Table 1 summarizes the demographics information of the included participants.

**Table 1.**
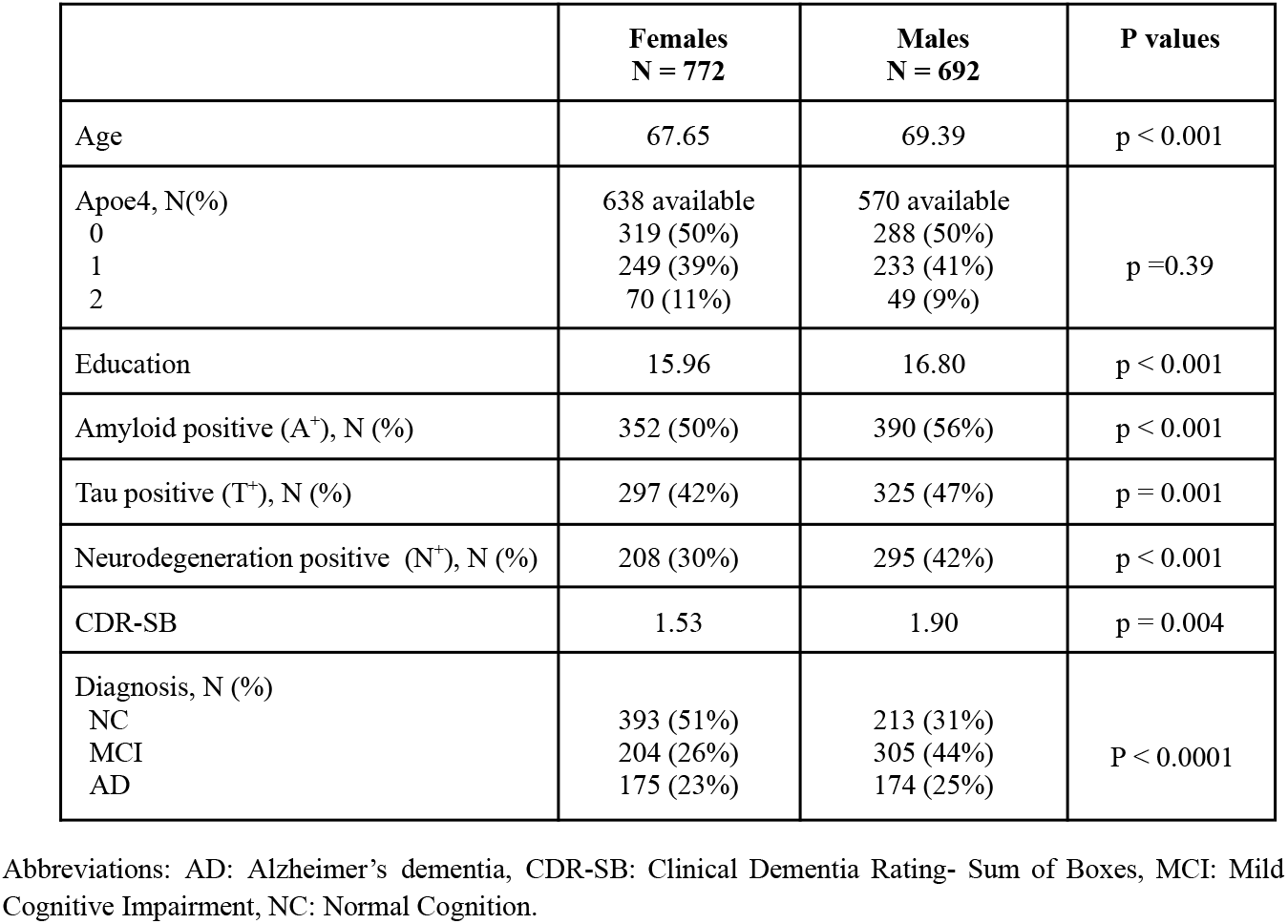
Baseline characteristics of the participants included in this study.

### Statistical Analyses

Baseline demographic characteristics were compared across sexes using independent sample t-tests for continuous measures and chi-square (χ^2^) tests for categorical measures. The following longitudinal mixed effects models were used to examine the differences in rates of cognitive decline across ATN groups (Model 1) as well as potential sex differences in the rates of decline within each ATN group (Model 2):

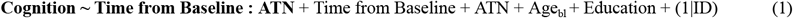

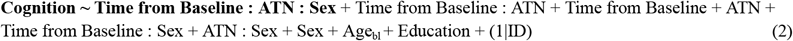

Where cognition indicates CDR-SB scores, time from baseline indicates the time between each longitudinal time point and the baseline clinical assessment visit, ATN indicates a categorical variable reflecting positivity on amyloid, tau, and neurodegeneration. Sex is a categorical variable contrasting males versus females. Age at baseline (Age_bl_) and Education were included as continuous fixed covariates in all models. For Model 1, the variable of interest was **Time from Baseline : ATN**, reflecting the potential differences in rates of cognitive decline across groups of interest.

For Model 2, the variable of interest was **Time from Baseline : ATN : Sex**, reflecting potential sex differences in rates of cognitive decline across the groups of interest. To ensure that potential differences in age (as the female participants were younger than the male participants in the full sample) and clinical diagnoses of the participants included (as there was a higher proportion of cognitively normal females compared to males in the full sample) did not impact the results, the analyses were repeated in an age- and diagnosis-matched subset of the dataset (N = 1146, 573 females, 573 males).

To assess whether there were potential differences in severity of pathologies between males and females (e.g. whether T^+^ females had greater burden of tau tangles than males), we further examined the sex differences in post-mortem amyloid and tau levels (using variables NPTHAL for Thal amyloid phase, NACCNEUR for CERAD amyloid score, and NACCBRAA for Braak tau stage) in the subset of the participants that had post-mortem pathology assessments available (N_Braak_: 68 males and 41 females; N_Thal_: 70 males and 42 females, N_CERAD_: 70 males and 42 females). The following ordinal logistic regression models were used for these analyses, adjusting for age at time of death.

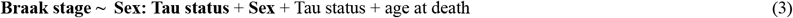

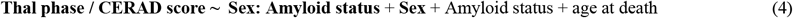

Where Amyloid and Tau status denote in vivo A and T status, and Braak, Thal, and CERAD denote the standard postmortem pathology staging scores of the same participants. The variables of interest included sex and sex interaction with amyloid or tau status.

Finally, we examined whether there are sex differences in the mediating effect of tau positivity on the associations between amyloid positivity and cognitive decline. We first confirmed the mediating relationships between amyloid, tau, and cognitive decline by performing a mediation analysis, with amyloid status as the independent variable, rate of cognitive decline as the dependent variable, and tau status as the mediating variable. Age and education were included as covariates. Rate of cognitive decline was estimated based on the random slopes of the following mixed effects models, similar to previous studies^23,24^ in the literature:

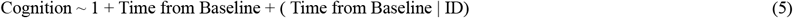

Sex was then examined as a potential moderator for this mediation effect. For both analyses, we used 1,000 bootstrapped resamples to estimate the confidence intervals. All statistical analyses were performed in R. The matched subsample was produced using MatchIt^25^ R package. lme4^26^ and lmerTest^27^ packages were used for mixed effects modelling and lavaan^28^ package was used for the mediation and moderation analyses.

## Results

Female participants were younger and had lower levels of education (p < 0.001). Furthermore, fewer females were A^+^, T^+^, or N^+^. Females also had lower CDR-SB scores at baseline (p = 0.004), and a greater proportion of them were classified as cognitively normal at the baseline visit assessment, while a greater proportion of males were classified in the mild cognitive impairment (MCI) group (p < 0.0001). As expected, there were no significant differences in age and clinical diagnoses of the age- and diagnosis matched subsample.

### Cognitive decline across different ATN groups

Figure 1 and Supplementary Table 1 summarize the estimated effects of ATN pathology on cognitive trajectories as measured by CDR-SB across time (Model 1). There was no significant impact of age at baseline, while education showed a small but significant impact (*β* = -0.1, p = 0.001). Except for the A^-^ T^+^ N^-^ group, all other included positive ATN pathology groups showed significantly higher baseline CDR-SB scores compared to the control group (A^-^ T^-^ N^-^). Differences were smaller for the A^+^ T^-^ N^-^ (*β* = 0.76, p = 0.02), followed by A^+^ T^-^ N^+^ (*β* = 1.87, p < 0.001), A^+^ T^+^ N^-^ (*β* = 2.03, p < 0.001), and A^+^ T^+^ N^+^ (*β* = 3.50, p < 0.001). Similarly, all pathology positive groups showed faster rates of cognitive decline compared to the reference A^-^ T^-^ N^-^. As expected, participants with only one pathology present showed less steep slopes of decline (A^+^ T^-^ N^-^: 0.1 increase per year, A^-^ T^+^ N^-^: 0.1 increase per year, and A^-^ T^-^ N^+^: 0.28 increase per year, all p values <0.005), while groups with multiple pathologies present showed even faster progression (A^+^ T^-^ N^+^: 0.75 increase per year, A^+^ T^+^ N^-^: 0.76 increase per year, and A^+^ T^+^ N^+^: 0.87 increase per year, all p values < 0.00001).

**Figure 1.**
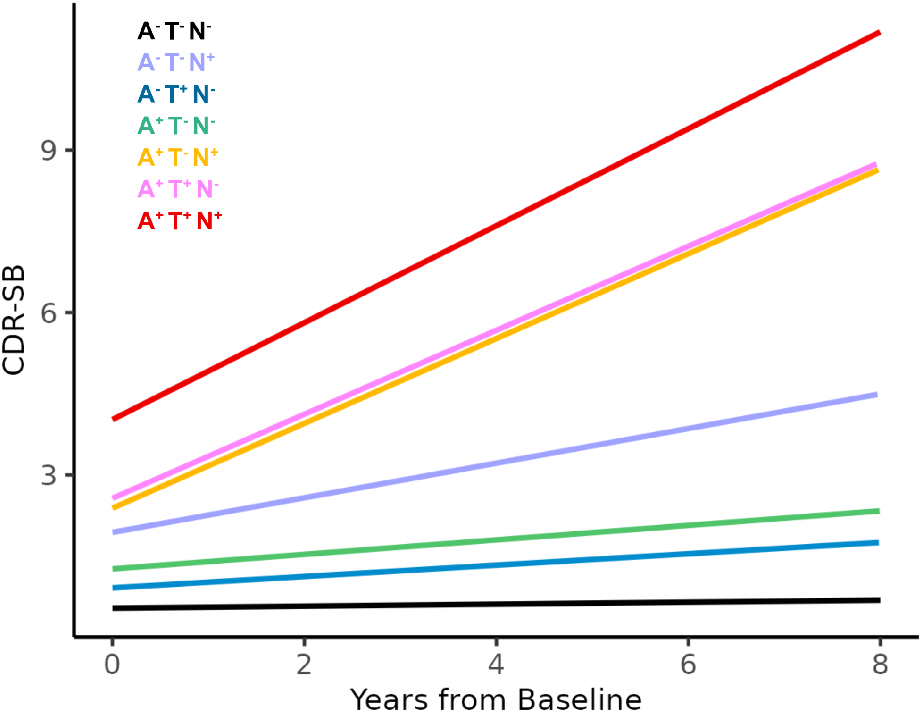
Longitudinal cognitive decline trajectories across ATN groups. A: Amyloid. T: Tau. N: Neurodegeneration. CDR-SB: Clinical Dementia Rating-Sum of Boxes.

### Sex differences in rates of cognitive decline across ATN groups

Figure 2 and Supplementary Table 2 summarize the estimated sex differences in the rates of cognitive decline across the ANT groups (Model 2). There were no significant sex differences in baseline CDR-SB scores across ATN groups. While there were no significant sex differences in the rates of cognitive decline in any of the A^-^ groups, we found a dichotomy based on tau status in A^+^ groups, with the A^+^ T^-^ N^-^ males showing an additional 0.17 increase in CDR-SB per year compared to A^+^ T^-^ N^-^ females, and the A^+^ T^-^ N^+^ males showing an additional 0.40 in CDR-SB per year compared to A^+^ T^-^ N^+^ females (p values < 0.005). Further still, female participants showed faster progression compared to males in both A^+^ T^+^ groups, with the A^+^ T^+^ N^-^ females showing an additional 0.24 increase in CDR-SB per year compared to A^+^ T^+^ N^-^ males, and the A^+^ T^+^ N^+^ females showing an additional 0.61 in CDR-SB per year compared to A^+^ T^+^ N^+^ males (p values < 0.00001). Repeating the analyses in the age- and diagnosis-matched subsample of the data yielded similar results in terms of significance and direction of the estimated effects (Supplementary Table 3).

**Figure 2.**
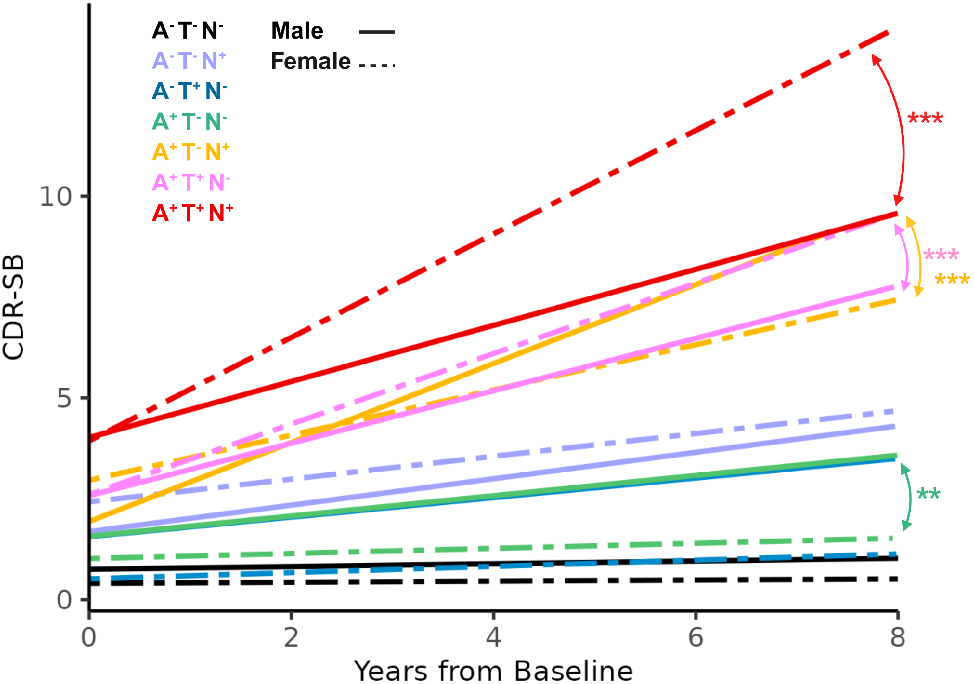
Sex differences in longitudinal cognitive decline trajectories across ATN groups. A: Amyloid. T: Tau. N: Neurodegeneration. CDR-SB: Clinical Dementia Rating-Sum of Boxes. Arrows indicate ATN groups that showed significant slope differences between males and females. ** p < 0. 005. *** p < 0.0001.

### The impact of amyloid on cognitive decline is mediated via tau and moderated by sex

Mediation analyses confirmed that tau positivity (T) mediated the association between amyloid positivity (A) and the rate of cognitive decline, indexed by the rate of increase in CDR-SB (Figure 3.A), while controlling for age at baseline and education. A was significantly associated with T (a = 0.720, *z* = 40.1, *p* < .001). In turn, T was positively associated with the rate of CDR-SB increase (b = 0.531, *z* = 9.02, *p* < .001). The direct effect of A on the rate of cognitive decline remained significant after accounting for the indirect path through T (c′ = 0.497, *z* = 9.22, *p* < .001), indicating a partial mediation. The indirect effect of A on the rate of cognitive decline via T was statistically significant (estimate = 0.382, 95% CI [0.298, 0.476], *p* < .001), as was the direct effect (0.497, 95% CI [0.391, 0.602], *p* < .001). The total effect of A on CDR-SB change was 0.879 (95% CI [0.798, 0.958]), with approximately 43.5% of the effect mediated by T (95% CI [34.1%, 53.9%]).

**Figure 3.**
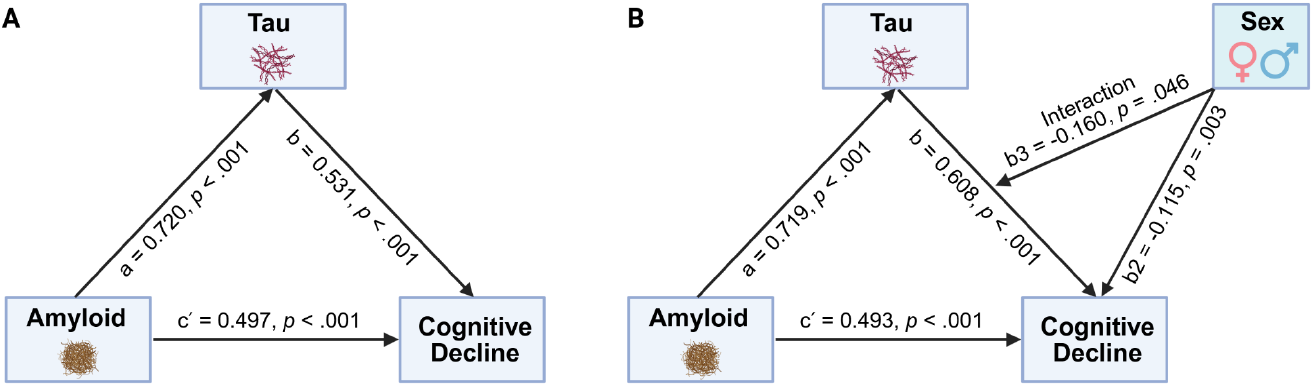
Sex differences in the mediating effect of tau positivity on the association between amyloid positivity and cognitive decline. A. Tau mediates the relationship between amyloid and cognitive decline. B. Sex moderates the mediation relationship between amyloid, tau, and cognitive decline. Cognitive decline indicates the rate of change in CDR-SB scores.

Furthermore, we found a marginal moderating effect of sex on this mediation, where females showed a stronger mediation effect compared to males (Figure 3.B). The main pattern of associations remained consistent with a significant association between A and T (a = 0.719, *z* = 39.4, *p* < .001), T and the rate of CDR-SB increase (b = 0.608, *z* = 8.56, *p* < .001) as well as direct effect of A on the rate of CDR-SB increase after accounting for the indirect path through T (c′ = 0.493, *z* = 9.32, *p* < .001). Sex had a significant main effect on the rate of cognitive decline (b2 = -0.115, *z* = -2.96, *p* = .003), with males showing a slower decline. The interaction between Tau status and sex suggested a trend toward a weaker effect of tau on decline among males (b_3_ = -0.160, *z* = -2.03, *p* = .045). The indirect effect of A on rate of CDR-SB increase was significant in both sexes, but larger in females (0.437, 95% CI [0.336, 0.544], *p* < .001) than in males (0.322, 95% CI [0.222, 0.422], *p* < .001) with the proportion of the total effect mediated by T being 47.0% in females (95% CI [37.2%, 56.5%]) compared to 39.5% in males (95% CI [28.1%, 50.5%]). The difference in indirect effects between sexes approached statistical significance (0.115, 95% CI [-0.008, 0.229], *p* = .046), suggesting possible moderating effect of sex on the mediation pathway.

### Sex differences in post-mortem amyloid and tau burden

Adjusting for age at time of death, as expected, T^+^ participants had significantly higher postmortem Braak tangle score (*β* = 2.35, p < 0.0001), and A^+^ participants had significantly higher postmortem amyloid burden as measured by both Thal phase (*β* = 1.23, p = 0.02) and CERAD neuritic plaque score (*β* = 3.07, p < 0.0001). There were no significant sex differences in postmortem amyloid and tau burden across any of the A, T, or N groups (all p values > 0.05).

## Discussion

This study examined the longitudinal cognitive decline trajectories of aging individuals on the spectrum of AD according to their ATN classifications, and whether there were any sex differences in these trajectories within each ATN group. Our findings showed differences in rates of cognitive decline across ATN groups, whereby groups that were positive on multiple biomarkers showed steeper decline. Our main finding revealed a specific dichotomic sex difference between tau positive versus tau negative biomarker groups that had positive amyloid biomarker status. Specifically, in the A^+^ T^-^ group, males showed a faster cognitive decline than females in contrast to the A^+^ T^+^ group, in which females showed a steeper cognitive decline. These results suggest differences in the vulnerability to tau pathology in participants with positive amyloid biomarkers. This may have important clinical implications for the development of sex-specific treatments and emphasize the importance of studying sex differences in AD.

While no sex differences were observed in A^-^ groups, significant sex differences were found in the A^+^ groups. Specifically, males showed a steeper longitudinal cognitive decline than females in the A^+^ T^-^ group. In contrast, A^+^ T^+^ females showed faster progression compared to A^+^ T^+^ males. Our results further confirm the presence of a mediating effect of tau pathology on the associations between amyloid positivity and cognitive decline, and suggest that these associations might differ between the sexes. Postmortem studies have identified sex differences in AD tau pathology, with a higher burden in females^29,30^, which might explain the faster progression of cognitive decline in A^+^ T^+^ females. Additionally, a large post-mortem study of 1453 participants from the Religious Orders Study and the Rush Memory and Aging Project reported both higher tau and marginally higher amyloid burden in females^31^, adjusting for age at the time of death. To test this hypothesis, we investigated the subset of participants that had postmortem amyloid and tau burden information available (7.5% of the sample) and did not find higher pathology burden in females or any other differences in amyloid or tau burden. This might be due to the smaller sample size with available pathological data or a ceiling effect within the tau positive group with most participants having neurofibrillary Braak stage V and VI. Alternatively, several other co-pathologies, including possible comorbid Lewy body or vascular disease not clinically detected might have an effect on observed sex differences. While the mechanism underlying increased rates of cognitive decline for the male participants in the A^+^ T^-^ group is not clear to us, this may suggest a higher cognitive reserve in females than in males in earlier disease stages with positive amyloid biomarker. Accordingly, previous studies reported a higher cognitive reserve in females^17,32,33^.

Our findings have important implications for disease diagnosis and management. Sex differences in the relationships between presence of amyloid and tau pathologies and cognitive decline might translate into sex differences in treatment efficacy when individuals receive amyloid clearing therapies. Since these treatments generally target patients in earlier disease stages, who would be more likely to be A^+^ T^-^, removal of amyloid might prevent cognitive decline more effectively in males than in females. This is also in line with the current evidence from recent clinical trials, where male participants benefited more from the treatment compared to their female counterparts^8,9^. Similarly, this may suggest that tau-targeting agents^34^ might be more effective for female participants.

While our study includes a large sample size with considerable longitudinal assessments allowing for robust estimation of longitudinal cognitive trajectories across ATN groups, we acknowledge certain limitations. First, amyloid and tau positivity were defined based on either PET or CSF assessments, leading to potential variability in the measurements. However, the literature suggests strong agreements between PET and CSF based assessments for both amyloid and tau pathologies^35–37^. We also found similarly high levels of agreement (88.2% for amyloid status and 87.4% for tau status) in the subsets of participants that had both CSF and PET data available, suggesting that the impact of using either biomarker would be minimal on the results in the context of this study. In addition, sex differences were found in the demographic characteristics of the included participants, whereby the female participants were younger, and the sample included a higher proportion of female participants that were cognitively normal at baseline. To control for these differences, the sex difference analyses (Model 2) were repeated in an age- and diagnosis-matched subset of the data, and the obtained results remained similar. Finally, due to data availability and sample size limitations, we were not able to incorporate vascular pathology (V) in the analyses. Since vascular pathology is a common comorbidity that exacerbates cognitive decline in aging individuals as well as AD patients^38–42^ and previous studies have reported sex differences in prevalence and impact of vascular pathology on cognitive decline trajectories^43,44^, future studies assessing sex differences in cognitive decline trajectories across ATN-V groups are warranted.

In conclusion, we identified a distinct sex-specific dichotomy in cognitive decline trajectories among individuals with amyloid positive biomarkers. Specifically, males exhibited a more rapid cognitive decline than females in the absence of tau pathology, whereas females experienced a steeper decline when tau biomarkers were also present. These findings may hold significant clinical implications for the development of sex-specific therapeutic strategies for AD.

## Data Availability

All data produced in the present study are available upon reasonable request to the authors.

https://naccdata.org/

## Author Contributions

All authors were involved with design, conceptualization, and interpretation of the findings. Yashar Zeighami and Mahsa Dadar completed the analyses and drafted the manuscript. All authors revised and approved the submitted version.

## Acknowledgements

The National Alzheimer’s Coordinating Center (NACC) database is funded by National Institute on Aging (NIA)/National Institutes of Health (NIH) Grant U24 AG072122. NACC data are contributed by the NIA-funded Alzheimer’s Disease Research Centers (ADRCs): P30 AG062429 (PI James Brewer, MD, PhD), P30 AG066468 (PI Oscar Lopez, MD), P30 AG062421 (PI Bradley Hyman, MD, PhD), P30 AG066509 (PI Thomas Grabowski, MD), P30 AG066514 (PI Mary Sano, PhD), P30 AG066530 (PI Helena Chui, MD), P30 AG066507 (PI Marilyn Albert, PhD), P30 AG066444 (PI David Holtzman, MD), P30 AG066518 (PI Lisa Silbert, MD, MCR), P30 AG066512 (PI Thomas Wisniewski, MD), P30 AG066462 (PI Scott Small, MD), P30 AG072979 (PI David Wolk, MD), P30 AG072972 (PI Charles DeCarli, MD), P30 AG072976 (PI Andrew Saykin, PsyD), P30 AG072975 (PI Julie A. Schneider, MD, MS), P30 AG072978 (PI Ann McKee, MD), P30 AG072977 (PI Robert Vassar, PhD), P30 AG066519 (PI Frank LaFerla, PhD), P30 AG062677 (PI Ronald Petersen, MD, PhD), P30 AG079280 (PI Jessica Langbaum, PhD), P30 AG062422 (PI Gil Rabinovici, MD), P30 AG066511 (PI Allan Levey, MD, PhD), P30 AG072946 (PI Linda Van Eldik, PhD), P30 AG062715 (PI Sanjay Asthana, MD, FRCP), P30 AG072973 (PI Russell Swerdlow, MD), P30 AG066506 (PI Glenn Smith, PhD, ABPP), P30 AG066508 (PI Stephen Strittmatter, MD, PhD), P30 AG066515 (PI Victor Henderson, MD, MS), P30 AG072947 (PI Suzanne Craft, PhD), P30 AG072931 (PI Henry Paulson, MD, PhD), P30 AG066546 (PI Sudha Seshadri, MD), P30 AG086401 (PI Erik Roberson, MD, PhD), P30 AG086404 (PI Gary Rosenberg, MD), P20 AG068082 (PI Angela Jefferson, PhD), P30 AG072958 (PI Heather Whitson, MD), and P30 AG072959 (PI James Leverenz, MD).

## Data Availability Statement

Data used in preparation of this article was obtained from the National Alzheimer’s Coordinating Center (NACC; https://naccdata.org/) database.

